# Quantification of patient motion in FDG-PET/CT brain imaging

**DOI:** 10.64898/2026.09.03.26362159

**Authors:** Kateřina Macková, Kateřina Dudášová, Petr Ježdík, Adam Kalina, Matyáš Ebel, Radek Janča

**Affiliations:** Department of Circuit Theory, Czech Technical University in Prague, Faculty of Electrical Engineering, Technicka 2, Prague 6, 160 00, Czech Republic; Department of Neurology, Second Faculty of Medicine, Charles University, Motol and Homolka University Hospital, a full member of European Reference Network EpiCARE, V Uvalu 84/1, Prague 5, 150 06, Czech Republic; Department of Paediatric Neurology, Second Faculty of Medicine, Charles University, Motol and Homolka University Hospital, a full member of European Reference Network EpiCARE, V Uvalu 84/1, Prague 5, 150 06, Czech Republic

**Keywords:** Brain, Epilepsy, Dynamic FDG-PET/CT, Head motion, Motion quantification

## Abstract

**Objectives:** FDG-PET is an important imaging tool in evaluation of neurological disorders, including drug-resistant epilepsy. Head motion can degrade PET image quality; however, the extent in routine examinations remains insufficiently characterized. This retrospective study aimed to quantify the magnitude and pattern of head motion in brain FDG-PET/CT and establish displacement-based scores for describing motion severity.

**Materials & Methods:** The analysis included 140 patients with focal drug-resistant epilepsy undergoing brain FDG-PET/CT, with 27 examined under general anesthesia as a reference group. PET acquisition comprised five consecutive 2-min frames. Head motion was quantified by rigid PET-to-PET registration and expressed as maximal displacement. Two experienced readers independently graded motion on a four-point ordinal scale. Ordinal logistic regression with repeated 10-fold cross-validation was used to derive displacement thresholds corresponding to visual motion scores.

**Results:** Patients without anesthesia demonstrated greater maximal displacement than the reference group (median, 2.6 vs. 0.7 mm; *p* < 0.001). Displacement increased throughout acquisition, peaking in the final frame in 85.7% of examinations. Motion predominantly involved inferior–superior translation and rotation around left–right axis, corresponding to head flexion–extension. Maximal displacement was strongly associated with visual motion grades (Kendall’s τ=0.68–0.70; *p* < 0.001). Derived thresholds classified displacement >3.25 mm as moderate and >6.4 mm as severe motion. In total, 40.7% of patients without anesthesia demonstrated moderate or severe motion, with severe motion in 12.4%.

**Conclusion:** Clinically relevant head motion was frequent in brain FDG-PET/CT despite head fixation, predominantly characterized by flexion–extension. Registration-based displacement provided an objective measure closely associated with expert visual assessment.

**Key points:**

- This study investigated the frequency of clinically relevant head motion in brain FDG-PET/CT and whether image-based displacement can provide an objective measure of motion severity.
- Clinically relevant head motion occurred in approximately 40% of brain FDG-PET/CTs, with severe motion in 12.4% of examinations and flexion–extension being the predominant pattern.
- Direct quantitative assessment of head motion from dynamic PET data may improve recognition of clinically relevant motion and support objective decisions on motion correction and repeat acquisition, potentially improving the reliability of brain FDG-PET/CT interpretation.

## 1. Introduction

Brain positron emission tomography (PET) is commonly used in neurological and neuro-oncological disorders [1] Particularly at early disease stages, pathological changes may manifest as subtle alterations in radiopharmaceutical uptake [2], making high image quality and spatial resolution essential for their detection and localization. Spatial resolution is inherently limited by the physical properties of PET systems and data processing [3]. Patient- and acquisition-related factors can further degrade image quality, resulting in effective spatial resolution typically worse than the approximate 3–5 mm point spread function specified for modern PET systems [4]. Despite standardized pre-acquisition protocols and patient positioning, head motion remains a critical source of image degradation, affecting both spatial resolution and radiotracer uptake quantification [5].

Head motion during brain PET causes image blurring which may lead to misinterpretation or render images unusable [6] and can distort radiotracer distribution by increasing apparent target size while reducing its apparent uptake [7]. These effects are particularly relevant for quantitative assessment [8–13] and MRI-based partial volume correction used to improve effective spatial resolution in focal epilepsy, Parkinson’s disease, and Alzheimer’s disease [5, 9–12]. Motion may also cause misregistration between emission PET and transmission CT or MRI, resulting in inaccurate attenuation correction [14].

Current guidelines recommend assessment and correction of motion-affected data where possible [1]. However, they provide limited guidance on how motion should be quantified and what degree should prompt correction, the exclusion of affected data, or repeat examination. Patient motion is routinely assessed visually using sinograms or reconstructed dynamic sequences, but slow movement of a few millimeters may be difficult to detect despite the potential effect on image processing and clinical evaluation [15–17]. Head motion has been systematically quantified only in limited cohorts using heterogeneous systems and methodologies. To our knowledge, patients undergoing brain FDG-PET/CT for epilepsy evaluation have not been specifically studied [17–21].

This study aimed to quantitatively characterize the magnitude and patterns of head motion in routine brain FDG-PET/CT. Understanding patient motion may support optimization of head fixation and development of motion detection, correction, and post-processing techniques. Relating objective motion quantification to subjective image assessment may also help identify displacement thresholds associated with perceptible motion and establish practical criteria for motion management and image quality control.

## 2. Materials and Methods

### 2.1. Clinical data

This retrospective study was approved by the Institutional Ethics Committee of Motol University Hospital (2022/06/15-EK-602.24/22). Informed consent was obtained from all patients or their legal guardians. Patients were diagnosed with focal drug-resistant epilepsy at the Motol and Homolka epilepsy centers between 2021 and 2025 and met the following criteria: (1) dynamic brain FDG-PET/CT acquired on a Siemens Biograph mCT or Vision 600 scanner; (2) structural 3T brain MRI acquired using an epilepsy protocol at submillimeter resolution; and (3) no history of brain surgery. The youngest pediatric and non-cooperative patients were examined under general anesthesia (GA) and were considered the reference group, assuming minimal motion during image acquisition. Details of the anesthesia protocol are provided in Supplementary File 1.

FDG was administered intravenously at 1.43 MBq/kg (100 MBq/70 kg) for Vision 600 and 2.86 MBq/kg (200 MBq/70 kg) for mCT after at least 6 h of fasting. Patients rested with eyes closed for 30–60 min in a quiet, dimly lit room during tracer uptake according to guidelines [22]. The head was immobilized using a head holder (08096815, Siemens Healthineers); a polyethylene foam Headrest Complete Set (07445740, Siemens Healthineers) was used in patients under GA. After a 2-min adaptation period to minimize initial motion, low-dose CT for attenuation correction was followed by five consecutive 2-min FDG-PET frames for motion assessment: dynamic PET.

Dynamic PET frames were reconstructed using OSEM3D with scanner-specific settings (Vision: four iterations, five subsets, with TOF; mCT: five iterations, 24 subsets, without TOF), with normalization, decay, randoms and dead-time corrections, a 4-mm Gaussian low-pass filter, and without attenuation correction. Voxel sizes were 2.84×2.84×3 mm for Vision and 3.18×3.18×3 mm for mCT. Dynamic PET was used for motion analysis, whereas clinical evaluation was based on summed PET data from a period without significant motion and reconstructed with different settings.

Structural MRI was acquired on a 3T Siemens MAGNETOM Vida and Philips Ingenia scanner using an epilepsy protocol [23] with submillimeter in-plane resolution, typically 0.5×0.5×1 mm. Only T1-weighted (T1w) images were used in this study.

### 2.2. Image processing

To track motion during PET acquisition, five 2-min frames of dynamic PET were acquired. The first frame (PET_1_) served as the assumed motion-free reference (fixed) image for subsequent rigid-body registration of later dynamic frames potentially containing motion-degraded data (PET_2_–PET_5_). Rigid-body registration was performed using the SPM12 toolbox for MATLAB (ver. 2024a) [24]. The resulting transformation matrices, denoted as **T**_moving→fixed_, were used for subsequent motion assessment, where the subscripts indicate the moving and fixed images, respectively. For the dynamic PET frames, the transformations were therefore denoted as **T**_2→1_–**T**_5→1_.

Although low-dose CT precedes PET acquisition and might intuitively serve as the initial reference for motion assessment, the low mutual information between noisy dynamic PET and CT images often results in registration failure [17]. Therefore, T1w MRI was used as an intermediate registration step. As direct PET-to-T1w registration may also be distorted by differences in neck or other soft-tissue displacement, a gray matter mask (GM) extracted from T1w images using FastSurfer (ver. 2.4.2) [25] was used instead (**T**_GM→1_). The original T1w image was used for CT registration (**T**_CT→T1w_). The final CT-to-PET_1_ registration resulted from transformation (**T**_CT→1_ = **T**_GM→1_ · **T**_CT→T1w_). The approximate position of the anterior commissure in MRI was set as the origin for all images instead of the default PET origin in the chest. The registration pipeline is illustrated in **Error! Reference source not found.**.

### 2.3. Quantification of patient movements during PET acquisition

Possible head motion was decomposed into translations along coordinate axes (left–right; posterior– anterior; inferior–superior) and rotations about these axes. Each co-registration transformation matrix was decomposed into three translation and three rotation components using the *spm_imatrix* function of the SPM12 toolbox. Absolute values were used to assess motion magnitude.

The displacement of brain structures due to motion depends on the distance from the center of rotation (anterior commissure) and the translation components and is typically greater in lateral regions. Thus, displacement (*d_m_*) was calculated for voxels (*X*(*i*), *Y*(*i*), *Z*(*i*)) representing the brain (*i* ∈ **GM**) as the maximum Euclidean distance from the initial position to the position after rigid transformation between dynamic PET frames:

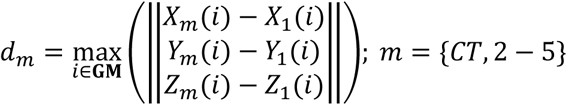

The maximal displacement relative to the first PET frame observed throughout the examination was defined as 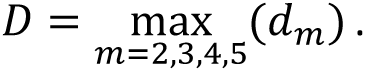 The quality of the final PET reconstructions depends on the number of detected counts individually influenced by motion. Thus, the average of displacement over frames was computed as 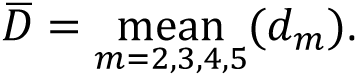

### 2.4. Subjective assessment of patient displacement in dynamic PET

Consistent with the routine clinical workflow at our centers, motion was visually assessed using cine playback of dynamic PET simultaneously in the axial, coronal, and sagittal planes, complemented by manual slice scrolling in 3D Slicer (www.slicer.org), see Supplementary File 2. Unregistered CT was displayed as a semi-transparent overlay on PET_1_.

Two experienced readers (RJ and KM) independently assessed all 140 examinations using a four-point ordinal scale: no motion – no conclusive visible shift; small motion – predominantly translational; moderate motion – translational and rotational components with displacements of a few millimeters; severe motion– displacements of tens of millimeters. CT-to-PET_1_ displacement was separately scored as no motion, inconclusive, or severe motion. Visual scores were compared with measured displacement and inter-reader agreement was assessed.

To quantify the association between maximal displacement and visual scores, ordinal logistic regression (*fitmnr*, MATLAB R2024a) was performed separately for each reader, detailed in Supplementary File 1. Displacement values corresponding to thresholds between visual scores were derived from the model and threshold uncertainties were estimated by nonparametric bootstrapping. Model performance on unseen data was assessed using repeated 10-fold cross-validation and summarized by exact agreement and proportions of one-, two-, and three-grade disagreements between predicted and reader-assigned grades.

### 2.5. Statistical analysis

Statistical analyses were performed in MATLAB R2024a (MathWorks, Natick, MA, USA). Significance was defined as *p* < 0.05. Associations between estimated head displacement and continuous variables (patient age, examination date) were assessed using Spearman’s rank correlation (ρ). Differences between groups (e.g., scanner model, anesthesia) were evaluated using the Mann–Whitney U test (U test) and Wilcoxon signed rank test for paired observations, with Cliff’s delta (δ) used to quantify effect size. The association between acquisition time and head displacement was assessed using a linear mixed-effects model (*fitlme*), detailed in Supplementary File 1.

Inter-reader agreement of the visual motion grading was assessed using weighted Cohen’s kappa (κ). The association between estimated head displacement and visual motion grading was evaluated using Kendall’s rank correlation (τ) and the Kruskal–Wallis test with post hoc pairwise comparisons.

## 3. Results

A total of 140 patients (70 women) met the selection criteria (median age: 15.9 years; interquartile range: 8.4–26.4; range: 1.2–64.2). Of 173 patients, 33 were excluded due to MRI (n=18), CT (n=5), or dynamic PET (n=8) unavailability and segmentation failure due to incomplete myelination (n=2). Of the 140 participants, 34 were examined with PET/CT Siemens Biograph mCT and 106 with Siemens Biograph Vision 600, of which 27 were under GA (**Error! Reference source not found.**). The dataset is summarized in Table 1 and presented in detail in Supplementary File 3.

**Table 1.**
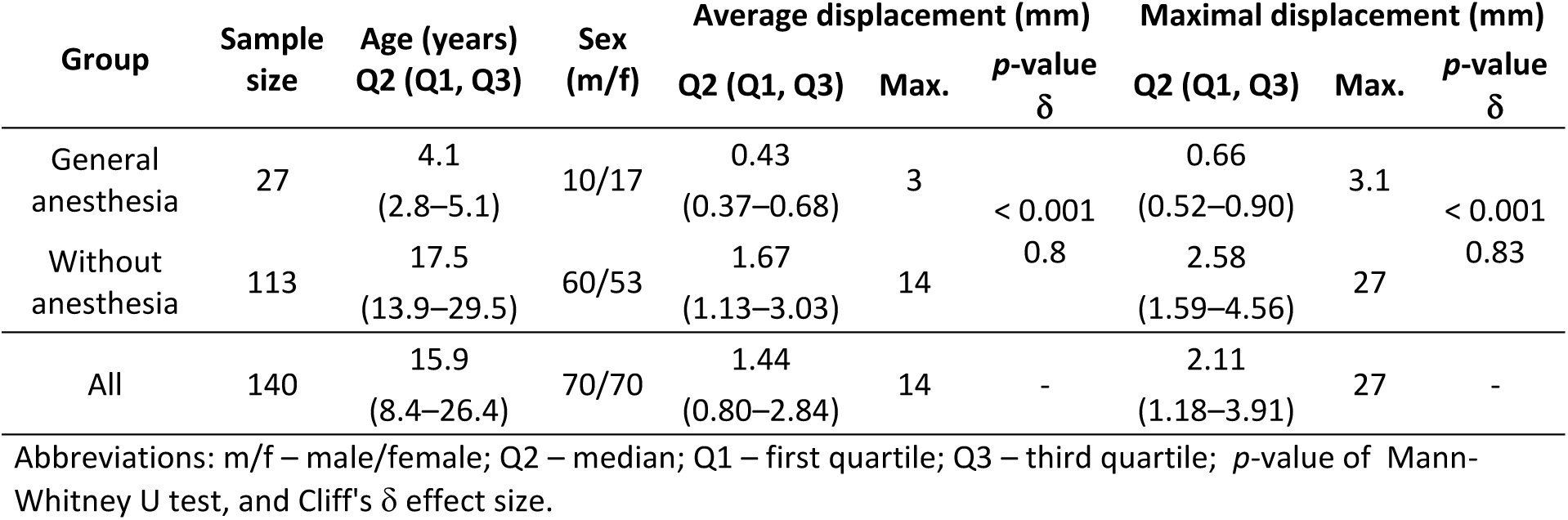
Patient characteristics and result of estimated displacement in dynamic PET.

### 3.1. Estimated displacement

Descriptive statistics are presented as median (interquartile range). The reference group comprised 27 patients under GA aged 4.1 (2.8–5.1) years (range: 1.2–13.3 years). The maximal displacement (*D*) was 0.7 (0.5–0.9) mm (max: 3.1 mm) and the average displacement (*D̄*) over frames was 0.4 (0.4–0.7) mm (max: 3 mm) (Table 1).

The 113 patients without GA, aged 17.5 (13.9–29.5) years, had a significantly higher maximal displacement of 2.6 (1.6–4.6) mm (max: 27 mm) and average displacement of 1.7 (1.1–3.0) mm (max: 14 mm) (*p* < 0.001, U test; δ>0.8) (**Error! Reference source not found.**b–c; Table 1). The maximal displacement was independent of the PET/CT scanner used (*p* = 0.13, U test), the examination date (ρ=−0.07, *p* = 0.38) and patient sex (*p* = 0.8, U test), see Supplementary File 1: Figures S1–3. The weak correlation ρ=−0.16 (*p* = 0.08) indicated higher displacement in younger patients although this did not attain statistical significance (**Error! Reference source not found.**a).

Displacement (*d_m_*) increased significantly across the 2-min frames in all translation and rotation axes, with acquisition time as a significant predictor in the linear mixed-effects model (all *p* < 0.001, see Supplementary File 1: Table S1). Largest displacement was observed in the last frame (PET_5_) in 85.7% of cases (120/140); this value was 7.9% (11/140) in PET_4_, 3.6% (5/140) in PET_3_, and 2.9% (4/140) in PET_2_. The dominant motion component corresponded to head flexion–extension compared with other directions: that is, absolute translation along the I–S axis and rotation about the R–L axis (*p* < 0.001, Kruskal–Wallis post hoc test) (**Error! Reference source not found.**). Directional motions are outlined in Supplementary File 1: Figure S4.

The displacement of CT related to PET_1_ was similar between groups with and without GA: 1.8 (1.1– 2.4) mm (max: 6.8 mm) and 1.9 (1.4–2.9) mm (max: 37 mm), respectively; (*p* = 0.26, U test). In the GA group, CT to PET_1_ registration via structural T1w MRI reduced the estimated displacement from 2.4 (1.8–3.6) mm with direct CT to PET_1_ registration to 1.8 (1.1–2.4) mm (*p* = 0.03, Wilcoxon signed rank test).

### 3.2. Subjective scored displacement

Visual inspection and subjective scoring of patient motion by RJ and KM reached substantial agreement weighted κ=0.64 for dynamic PET sequences. Correlation between maximal displacement and reader categories was τ=0.7 (*p* < 0.001) for RJ and τ=0.68 (p < 0.001) for MK. The scores categorized displacement (*p* < 0.001, Kruskal–Wallis test), except the post hoc difference between moderate and severe motion scores (*p* > 0.1, Kruskal–Wallis post hoc test). The maximal displacement scored as no motion by both readers was 0.7 (0.6–1.2) mm (41×); this value was 1.9 (1.5–2.1) mm (20×) for small, 4.1 (3.6– 4.8) mm (16×) for moderate, and 7.4 (5.4–9.6) mm (9×) for severe motion (**Error! Reference source not found.**).

Maximal displacement was strongly associated with visual scores for both readers based on ordinal logistic regression (KM: likelihood-ratio χ²=161.1, *p* < 0.001, R²=0.42; RJ: likelihood-ratio χ²=144.5, *p* < 0.001, R²=0.41). The estimated thresholds separating scores are summarized in Table 2. The model showed comparable agreement with visual scores for both readers in repeated 10-fold cross-validation. For KM, exact agreement was achieved in 66.1±1.2% cases, with one-grade disagreement in 31.7±1.2% and disagreement of two or more grades in 2.2±0.1% of cases; corresponding values for RJ were 68.5±1.1%, 30.8±1.1%, and 0.7±0%, respectively. Displacement showed a continuous distribution with overlap between adjacent grades, with no single set of strict boundaries evident. To obtain a common quantitative scoring, both readers’ thresholds were averaged, resulting in boundaries of 1.7–3.25 mm for small motion, 3.25–6.4 mm for moderate motion, and >6.4 mm for severe motion. Using these boundaries, 29.2% of patients (33/113) were classified as having no motion, 30.1% (34/113) as small motion, 28.3% (32/113) as moderate motion, and 12.4% (14/113) as severe motion.

**Table 2.**
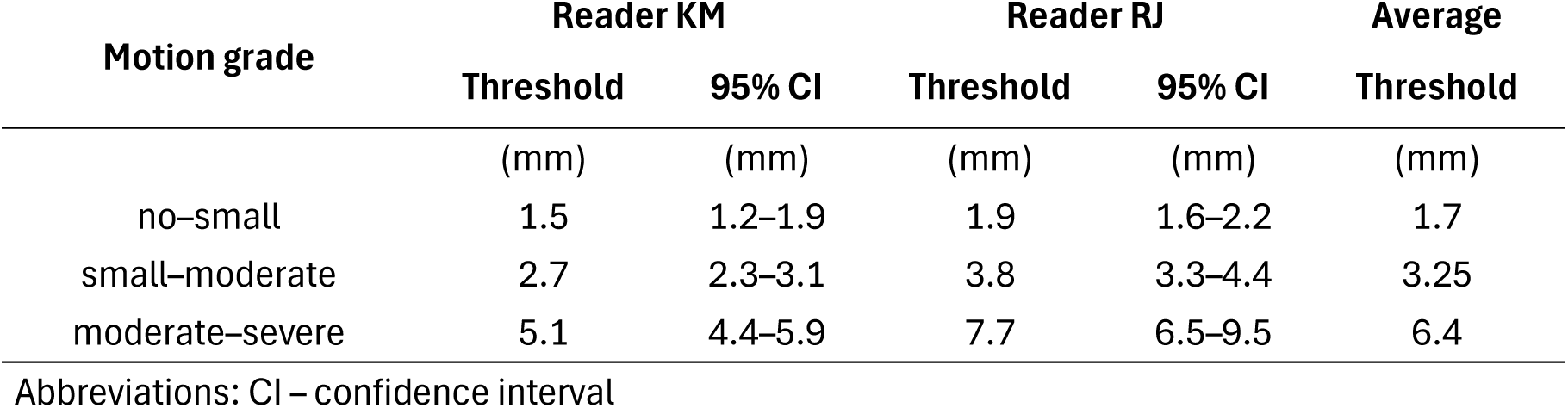
Result of ordinal logistic regression.

Visual inspection of CT to PET_1_ motion showed lower inter-reader agreement of weighted κ=0.24, with substantial overlap between visual scores (see Supplementary File 1: Table S2, Figure S5). The number of examinations assigned inconclusive and severe scores was insufficient for reliable ordinal regression modeling. Therefore, substantial CT to PET_1_ motion was defined using the Tukey’s fence for outliers *q*_0.75_ + 1.5 × (*q*_0.75_ − *q*_0.25_), yielding a 4.9-mm threshold. By this criterion, 8.9% of patients (10/113) had substantial motion. Details are provided in Supplementary file 2.

## 4. Discussion

This analysis of a cohort of patients with epilepsy undergoing brain FDG-PET/CT without GA revealed clinically relevant motion despite standardized positioning and head fixation [1, 2]. Motion was generally gradual, increasing throughout acquisition and peaking in the final dynamic frame in most patients. The predominant pattern was inferior–superior translation and rotation around the left–right axis, corresponding to nodding-type motion (head flexion–extension). Image-based displacement was strongly associated with subjective visual motion scores. Displacement >3.25 mm was classified as moderate motion and >6.4 mm as severe motion. Overall, 40.7% of patients exhibited moderate or severe motion, including 12.4% with severe motion.

The observed magnitude of motion is broadly consistent with that reported in previous studies, ranging from <1 mm in well-controlled examinations to >10 mm in individual cases. However, direct comparisons are limited by differences in radiotracers used, acquisition duration, reconstruction methods, and approaches to motion quantification [16, 17, 19–21, 26, 27]. Severe motion was observed in 12.4% of this cohort, comparable to the 12.6% reported by Kang et al. [18] and intraframe motion >3 mm in approximately 15% reported by Jin et al. [16]. Spangler-Bickell et al. reported high motion in 24% of examinations and minimal motion (≤1 mm) in 30%, similar to the 29% no-motion cases in our cohort; Spangler-Bickell et al. also reported an impact of motion correction on diagnostic image interpretation in 8% of cases [17]. Consistent with previous studies, the predominant motion pattern was characterized by inferior–superior translation and rotation around the left–right axis [13, 19, 21, 27].

This study has some limitations. First, head displacement was estimated using PET-to-PET image registration and is therefore subject to registration error. In the reference group (with GA), displacement was <1 mm in most examinations, suggesting a registration error of approximately ≤1 mm, consistent with previously reported registration accuracy [28]. Direct PET-to-CT registration was unreliable [17]; therefore, MRI was used as an intermediate reference, improving registration performance. However, cross-modality registration may introduce errors of approximately 1–2 mm [29–31]. Consequently, PETto-CT displacement within this range cannot be distinguished from registration uncertainty, whereas outliers exceeding 4.9 mm were identified in almost 9% of cases. Second, maximal displacement is a summary measure and does not directly quantify the effect of motion on image quality, which also depends on timing and the proportion of acquired counts affected [15–17]. We therefore also considered average displacement. However, future studies should directly relate quantitative motion to image quality and diagnostic performance for specific clinical indications. Third, visual motion scoring is inherently subjective, as reflected by the substantial but imperfect inter-reader agreement. The overlap in displacement between adjacent grades further indicates that motion is continuous rather than represented by distinct classes. The reported thresholds should therefore be interpreted as model-derived descriptors of expert visual grading and not definitive clinical acceptance limits. Clinically meaningful thresholds require studies linking quantitative motion to objective image quality and diagnostic accuracy, as previously demonstrated for the impact of motion correction on diagnostic interpretation [17]. Fourth, the temporal resolution of motion assessment was limited by the 2-min PET frames, preventing characterization of intraframe motion. While shorter frames would provide more detailed motion trajectories, they require specialized reconstruction and registration methods which are often vendor- or scanner-specific [17, 18, 32]. The use of 2-min frames falls within the 1–5-min range recommended by SNMMI/EANM for assessing motion in dynamic PET [1] and represents a practical approach using routinely acquired PET data and widely available registration software.

In conclusion, substantial head motion occurred in a clinically relevant proportion of patients undergoing brain FDG-PET/CT despite standardized positioning and head fixation, with over one in 10 examinations showing maximal displacement exceeding 6.4 mm, typically in the frontal and occipital poles. Predominant nodding-type motion identifies head flexion–extension as a potential target for improving fixation and reducing patient discomfort. Registration of dynamic PET frames provided an objective measure of head displacement closely reflecting expert visual assessment and could be derived directly from PET data without external motion-tracking equipment. This approach could complement subjective grading and improve motion assessment in routine brain PET. Further studies relating motion to image quality and diagnostic performance are needed to establish clinically relevant thresholds for guiding motion management, including motion correction, exclusion of degraded data, and repeat ac-quisition.

## Data Availability

All parametrized data and materials are available in the supplementary files. Access to the original clinical data may be granted on a case-by-case basis following individual review where an appropriate data sharing agreement has been established between the requesting institution and the data-owning institution(s).

## Abbreviations

CT: computed tomography
FDG: fluorodeoxyglucose
GA: general anesthesia
GM: gray matter
MRI: magnetic resonance imaging
OSEM3D: three-dimensional ordered-subsets expectation maximization
PET: positron emission tomography
PET/CT: positron emission tomography/computed tomography
PET/MR: positron emission tomography/magnetic resonance
T1w: T1-weighted

## Declarations

This study was approved by the institutional ethical committee of Motol University Hospital (2022/06/15-EK-602.24/22) in accordance with the Declaration of Helsinki of the World Medical Association and the International Ethical Guidelines for Biomedical Research Involving Human Subjects, prepared by the Council for International Organizations of Medical Sciences in collaboration with the World Health Organization, issued in Geneva, 1993. The data used was collected with the informed consent of all patients or their legal guardians.

## Consent for publication

Not applicable.

## Competing interests

The authors declare that they have no competing interests.

## Funding

RJ is a recipient of grants from the Czech Health Research Council (NU23-08-00528 Automatic detection and objective parametrization of hypometabolism in PET brain imaging). KM, KD, PJ, OB, AK are supported by the grant. RJ and PJ are supported by ERDF-Project Brain Dynamics (No. CZ.02.01.01/00/22_008/0004643). KM is supported by the grant agency of the Czech Technical University in Prague (SGS26/071/OHK3/1T/13). Additional support of this work was provided by the Ministry of Education, Youth and Sports of the Czech Republic through project LX22NPO5107 (NEUR-in): Financed by European Union – Next Generation EU.

## Author contributions

KM conceptualized the research goals (equal contribution), developed the methodology, performed the formal analysis and validation, wrote the original draft (equal contribution), and visualized the results (equal contribution). KD contributed to the methodology design and reviewed and edited the manuscript. PJ collected and curated the data. OB provided resources for clinical data collection and reviewed and edited the manuscript. AK and ME provided resources for clinical data collection. RJ conceptualized the research goals (equal contribution), contributed to the methodology and formal analysis, wrote the original draft (equal contribution), visualized the results (equal contribution), supervised the research, and acquired funding.

## Acknowledgements

The authors thank the staff at the participating clinics of Motol and Homolka University Hospital for their support and cooperation throughout this study: the PET Centre – Ass. prof. Otakar Belohlavek, Csc., Jiri Ters; the Department of Neurology – Prof. Petr Marusic, MD, PhD; David Krysl, MD, PhD; the Department of Pediatric Neurology – Prof. Pavel Krsek, MD, PhD; Martin Kudr, MD, PhD; Alena Jahodova, MD, PhD; Anezka Hejbalova (Belohlavkova), MD, PhD.Access to CESNET storage facilities provided by the project „e-INFRA CZ” under the programme „Projects of Large Research, Development, and Innovations Infrastructures” (LM2023054), is acknowledged. ChatGPT (OpenAI) was used to improve the grammar and style of the manuscript. ChatGPT and Gemini (Google) were also used to assist in identifying potentially relevant literature. All AI-assisted content was reviewed and verified by the authors, who take full responsibility for the final content of the manuscript.

---

**Figure 1.**
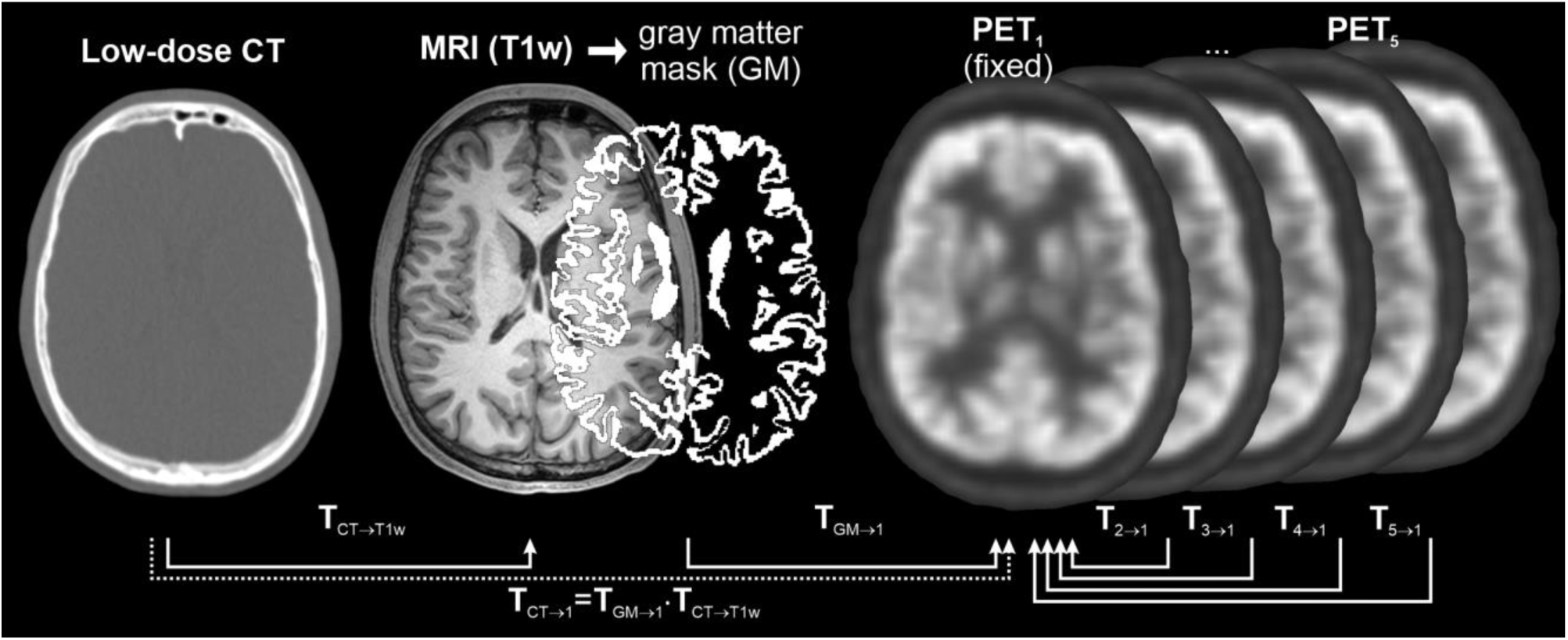
Scheme of registration pipeline. The first frame of dynamic PET_1_ was selected as a referential (fixed) image for registration. Other blocks (PET_2_–PET_5_) were rigidly registered to PET_1_ using obtained transformations **T**_2→1_ − **T**_5→1_. For MRI registration, a gray matter mask (GM) was extracted and registered to PET_1_ by transform **T**_GM→1_. CT was registered to PET_1_ using initial registration to structural MRI (GM to PET_1_ realignment, **T**_CT→1_ = **T**_GM→1_ · **T**_CT→T1w_). Decomposition of transformations to translation and rotation components resulted in displacement estimation in all axes.

**Figure 2.**
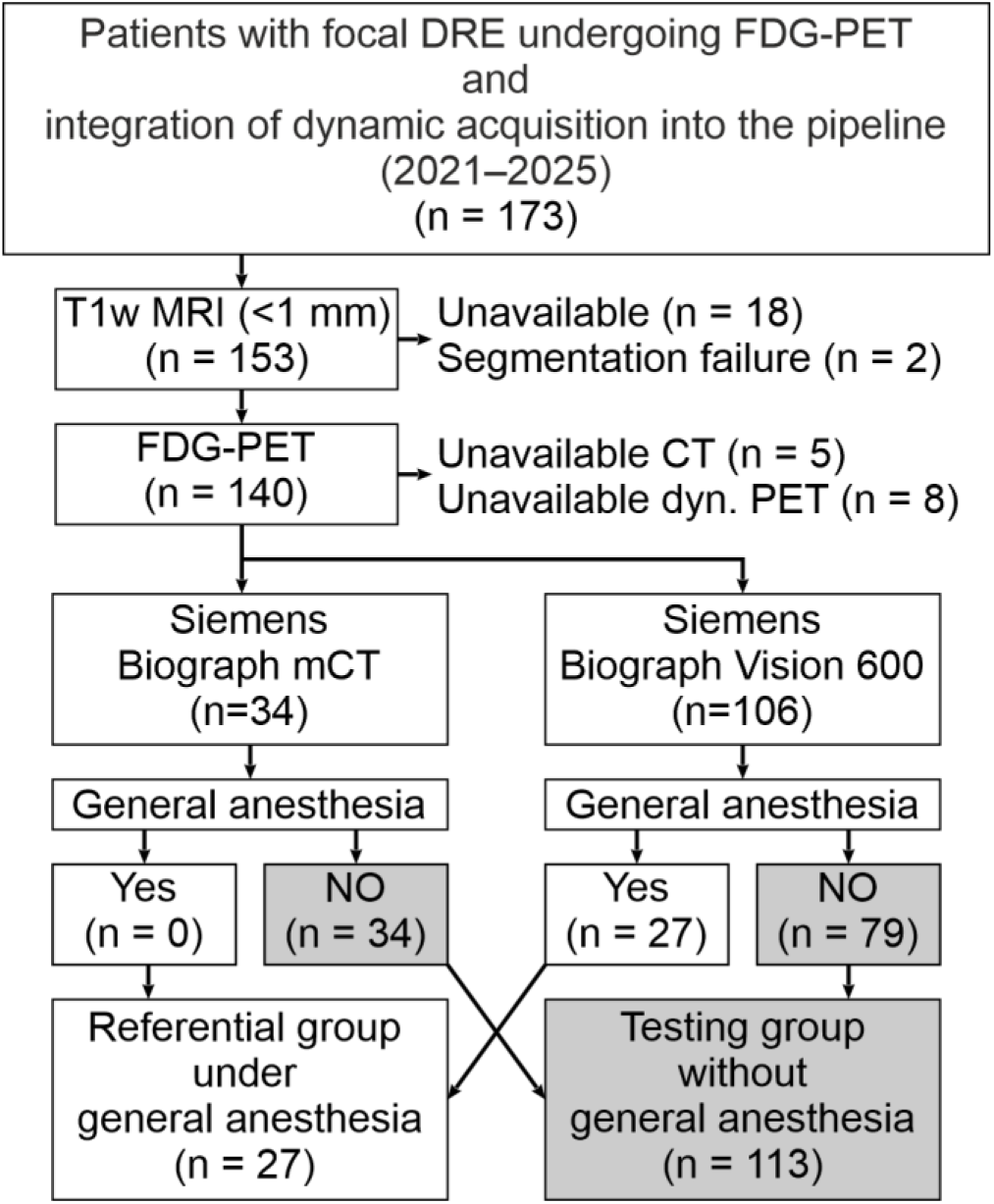
Flowchart of the selection of patients with focal drug-resistant epilepsy (DRE). Abbreviations: T1 weighted MRI – T1w.

**Figure 3.**
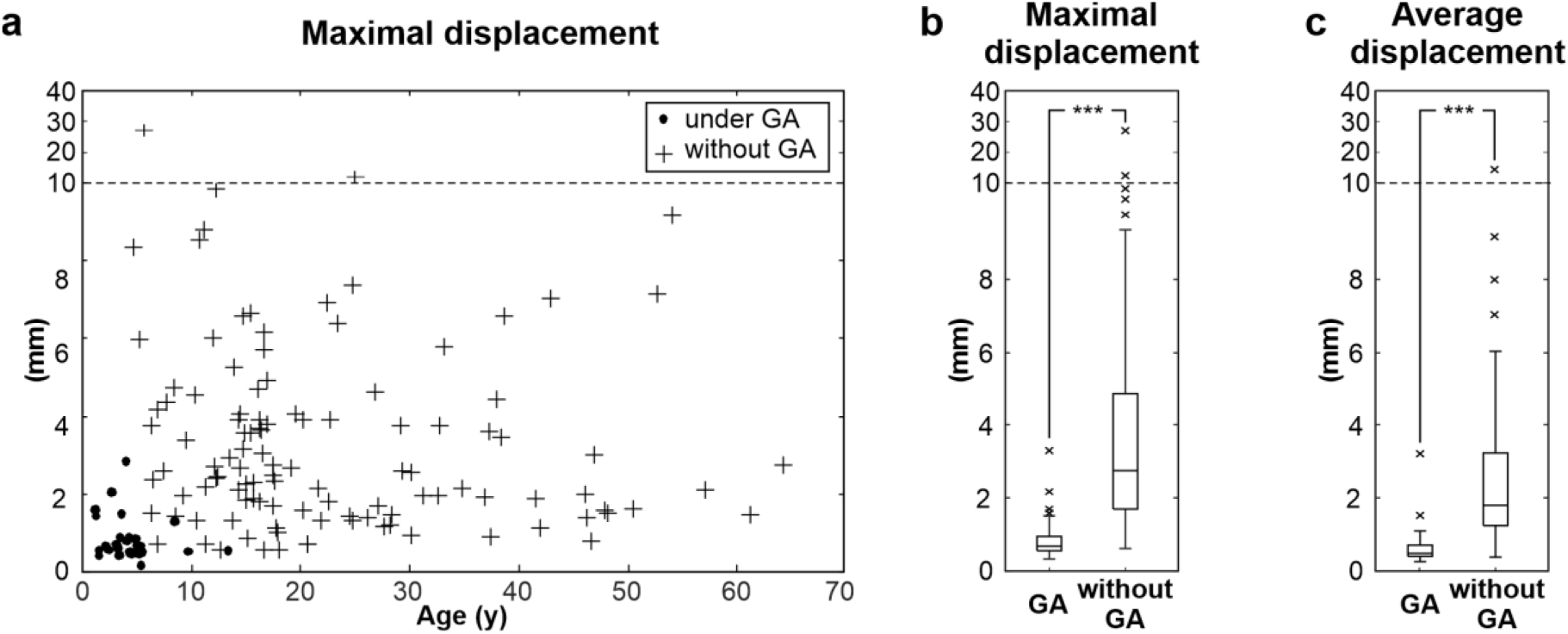
Quantified displacement: (a) maximal displacement was not significantly associated with patient age. Patients without general anesthesia (GA) exhibited higher motion quantified by (b) maximal displacement and (c) average displacement than those under GA (*p* < 0.001, U test). Note: the outlier part of the vertical axis continues in different linear scales.

**Figure 4.**
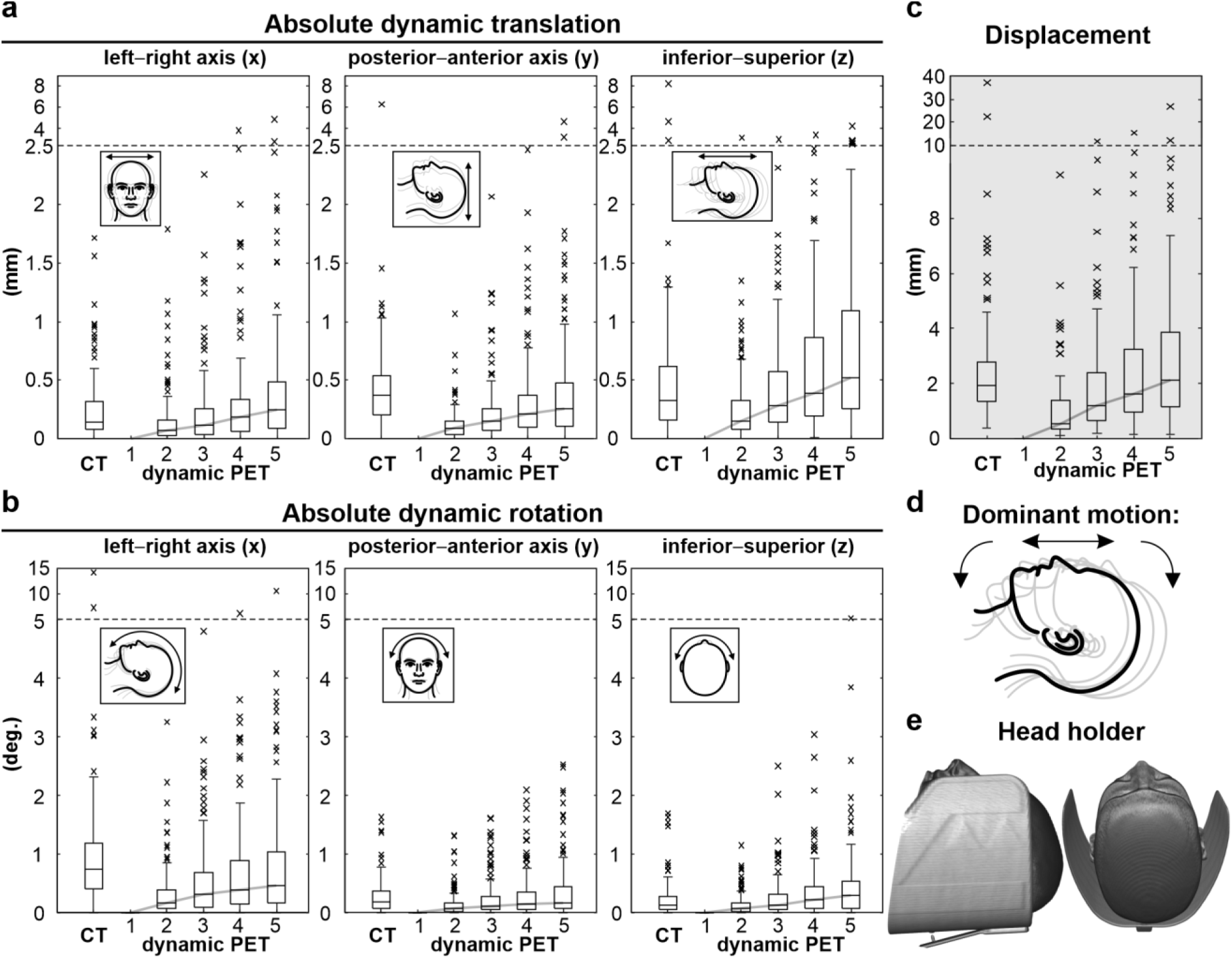
Estimated motion during dynamic PET acquisition: (a) absolute translation components; (b) absolute rotation components; (c) displacement (*d_m_*) at the most displaced part of the brain; (d) dominant motion following head flexion or extension; (e) 3D reconstruction of the head in the holder. Note: the outlier part of the vertical axis continues in different linear scales.

**Figure 5.**
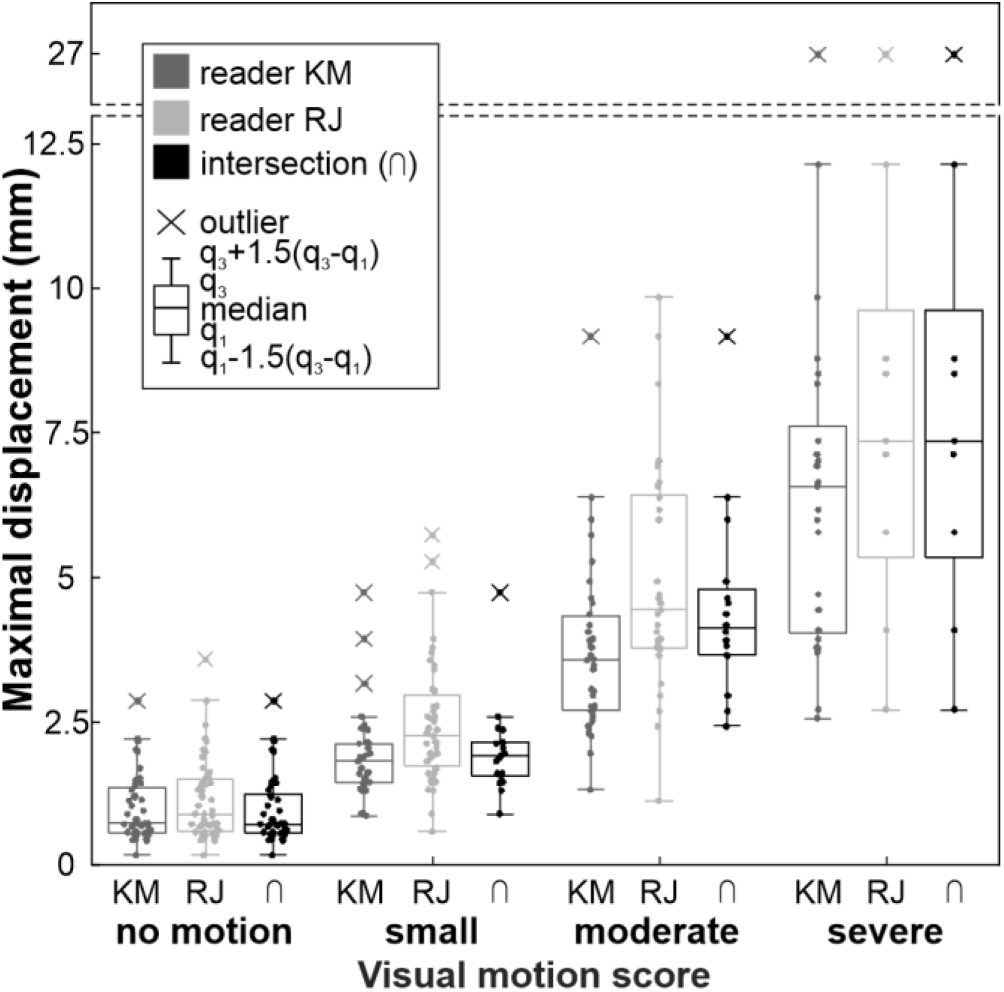
Scoring of dynamic PET maximal displacement by visual subjective assessment. The intersection (⋂) represents a match between both readers. Note: the outlier part of the y-axis is gapped; the upper part of the y-axis is omitted, and outliers are shown above the gap.

## Notes

### Competing Interest Statement

The authors have declared no competing interest.

