## Supplementary File 1 for "Quantification of patient motion in FDG-PET/CT brain imaging"

### 1. Supplementary Methods

#### 1.1. General Anesthesia Description

The youngest paediatric and non-cooperative patients underwent examination under general anaesthesia (GA), usually induced with intravenous propofol (2mg/kg) or midazolam and when required, maintained with sevoflurane at 0.6–0.8 age-adjusted minimum alveolar concentration via a face or laryngeal mask. Several commercially available anesthetic agents containing the same active substances are routinely used at the epilepsy centre.

#### 1.2. Ordinal logistic regression model

Ordinal logistic regression was performed separately for each reader using maximal head displacement as the predictor and the four-level visual motion score as the ordinal outcome. The model was fitted using the MATLAB *fitmnr* function (MATLAB R2024a).

Displacement values corresponding to decision thresholds between adjacent visual scores were estimated from the fitted models over a grid of 10 000 displacement values spanning the observed range. Uncertainty of the estimated displacement thresholds was assessed using nonparametric bootstrap resampling. For each reader, 1000 bootstrap samples of equal size to the original dataset were generated by sampling patients with replacement. The ordinal logistic regression model was refitted to each bootstrap sample, and the three decision thresholds were re-estimated. The 95% confidence intervals were defined by the 2.5th and 97.5th percentiles of the resulting bootstrap threshold distributions. Failed model fits were excluded from the corresponding bootstrap estimate.

The ability of maximal displacement to reproduce the reader-assigned motion grades was evaluated using 100 repetitions of 10-fold cross-validation. In each repetition, the dataset was randomly divided into 10 folds. The ordinal logistic regression model was fitted using nine folds and used to predict the motion grades in the remaining fold; this process was repeated until each examination had been evaluated. Agreement between predicted and reader-assigned grades was summarized as exact agreement and one-, two-, and three-grade disagreement, as well as Cohen’s weighted κ. Results were averaged across the 100 repetitions and expressed as mean ± standard deviation.

#### 1.3. Linear mixed effect model

To assess whether head motion increased with time spent in the scanner, linear mixed-effects models were fitted separately for each motion parameter using the MATLAB *fitlme* function (MATLAB R2024a). Absolute motion values were used for both translation and rotation parameters. Time point (dynamic PET frame) was included as a continuous fixed effect, with patient included as a random intercept to account for repeated measurements within patients. The significance of the time effect was assessed from the fitted model.

### 2. Supplementary Results

#### 2.1. CT–PET motion

Ordinal logistic regression was initially evaluated to derive displacement-based decision thresholds for the CT–PET_1_ visual motion grades. However, the number of examinations in the inconclusive and severe categories was low. During bootstrap resampling and cross-validation, some resampled or training datasets lacked one or more of the visual grades, resulting in failed ordinal model fits. Consequently, reliable decision thresholds and internally validated classification performance could not be obtained.

Because reliable ordinal modelling was not possible, substantial CT–PET_1_ motion was defined using the Tukey-fence criterion for outliers. This yielded a displacement threshold of 4.9 mm, above which examinations were classified as having substantial CT–PET_1_ motion. By this criterion, 10/113 (8.9%) patients showed substantial motion, with displacement of 7 (5.7– 8.9) mm, max. 37 mm, and 6.8 mm in a case (1/27) under GA.

Table S1. Linear mixed-effects model results for the association between head motion and acquisition time. For each motion parameter, a separate linear mixed-effects model was fitted with absolute motion magnitude as the dependent variable, dynamic PET frame (2-min intervals) as a continuous fixed effect, and patient as a random intercept. β represents the estimated change in absolute motion per 2-min frame. Confidence intervals are 95%. Translation parameters are reported in mm/2 min and rotation parameters in degrees/2 min.

| Motion parameter | β | lower 95% CI | upper 95% CI | units of β | p-value |
| --- | --- | --- | --- | --- | --- |
| Shift X | 0.09 | 0.07 | 0.11 | mm/2 min | <0.001 |
| Shift Y | 0.10 | 0.08 | 0.12 | mm/2 min | <0.001 |
| Shift Z | 0.17 | 0.15 | 0.19 | mm/2 min | <0.001 |
| Rot X | 0.19 | 0.15 | 0.22 | deg/2 min | <0.001 |
| Rot Y | 0.07 | 0.06 | 0.09 | deg/2 min | <0.001 |
| Rot Z | 0.11 | 0.09 | 0.13 | deg/2 min | <0.001 |
| 3D shift | 0.25 | 0.22 | 0.27 | mm/2 min | <0.001 |
| 3D rot | 0.26 | 0.22 | 0.30 | deg/2 min | <0.001 |
| Displacement | 0.64 | 0.56 | 0.73 | mm/2 min | <0.001 |

Table S2. Results of visual assessment of motion between the attenuation CT and first PET frame.

| Motion grade | Reader KM | | Reader RJ | | Both readers agreement | |
| --- | --- | --- | --- | --- | --- | --- |
|  | n | Displacement (mm) Q2(Q1–Q3) | n | Displacement (mm) Q2(Q1–Q3) | n | Displacement (mm) Q2(Q1–Q3) |
| No motion | 104 | 1.9 (1.3–2.5) | 135 | 1.9 (1.3–2.7) | 104 | 1.9 (1.3–2.5) |
| Inconclusive | 29 | 2.6 (1.5–3.8) | 3 | 5.9 (4.1–6.7) | 1 | 5.9 |
| Severe | 7 | 3.7 (2.3–18.6) | 2 | 29.7 (22.5–36.9) | 2 | 29.7 (22.5–37) |

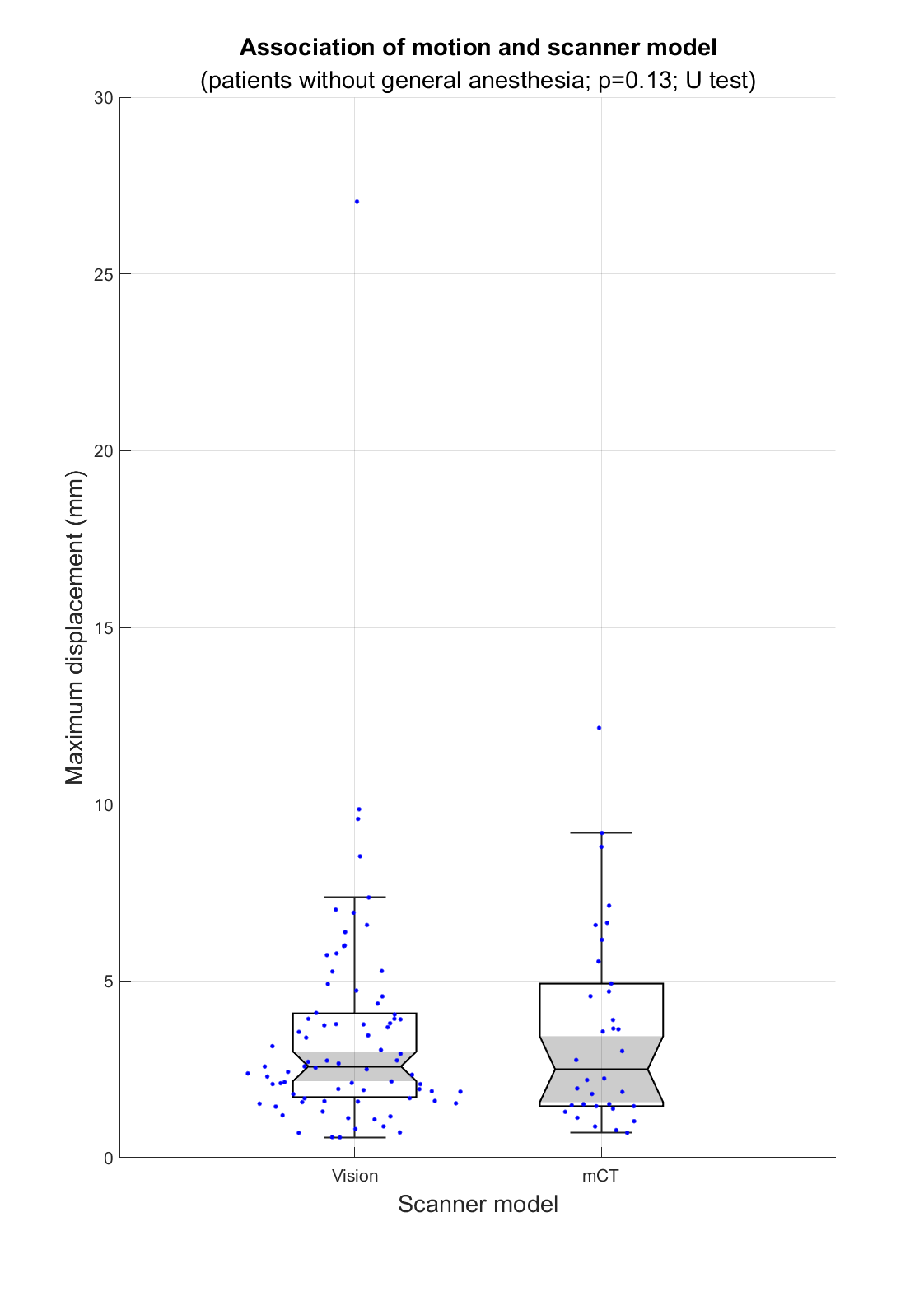

Figure S1. Comparison of maximal displacement between scanner models. Maximal displacement during the dynamic PET acquisition is shown for examinations performed on mCT and Vision 600 scanner models. Individual examinations are shown as points; box plots indicate the median and interquartile range.

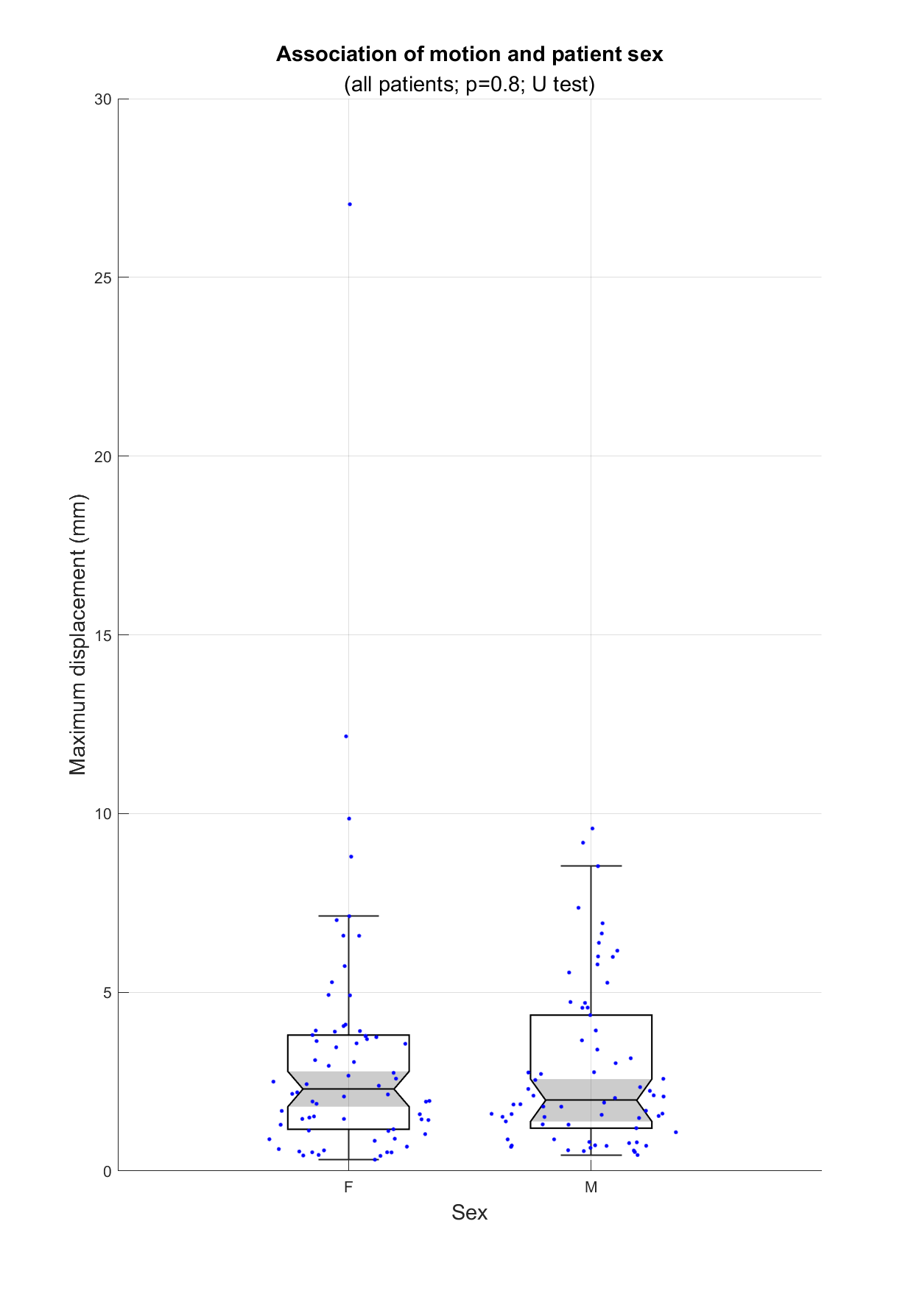

Figure S2. Comparison of maximal displacement by patient sex. Maximal displacement during the dynamic PET acquisition is shown for female (F) and male (M) patients. Individual examinations are shown as points; box plots indicate the median and interquartile range.

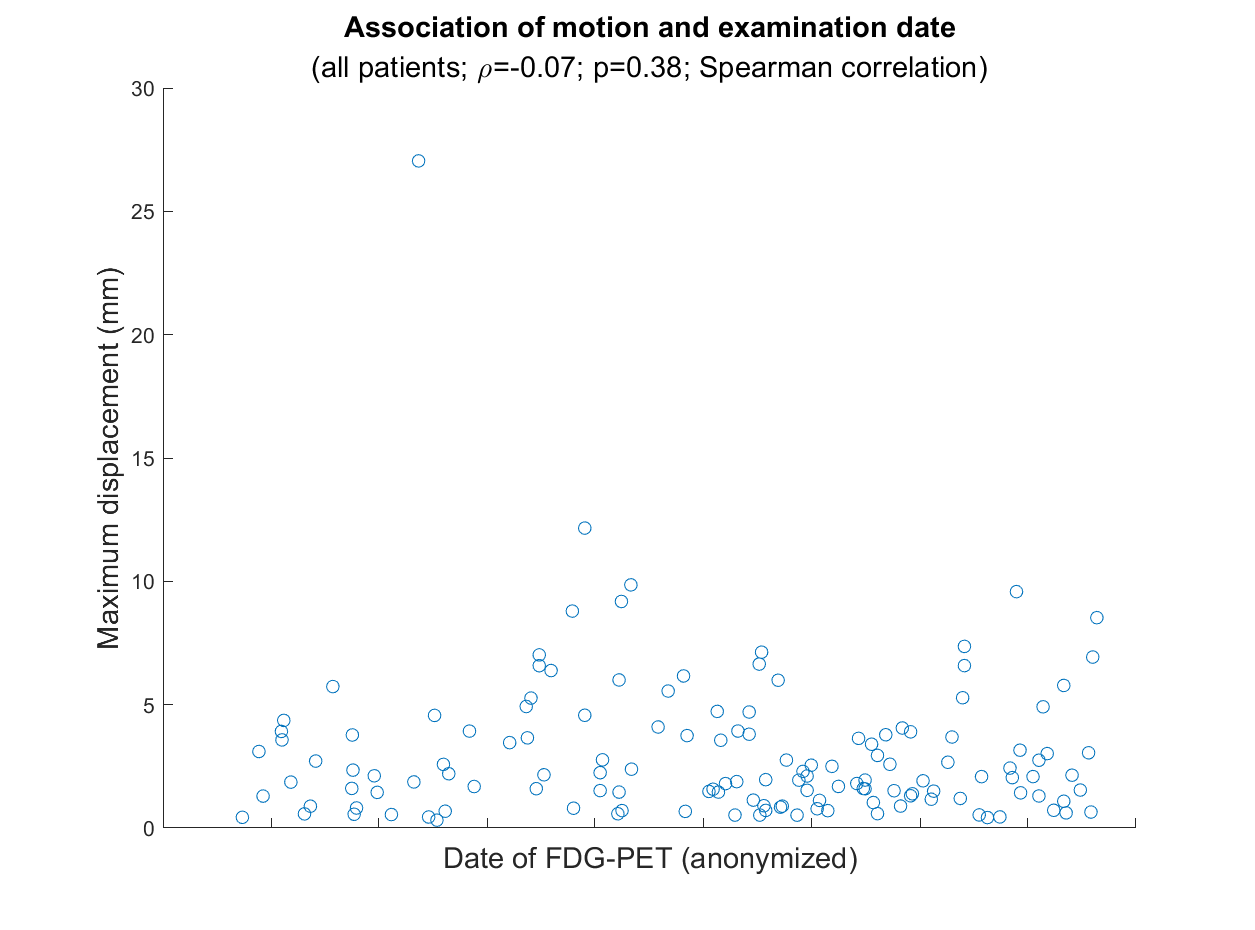

Figure S3. Association between maximal displacement and examination date. Maximal displacement during the dynamic PET acquisition is shown as a function of examination date. Each point represents one examination; the correlation was assessed using Spearman’s rank correlation.

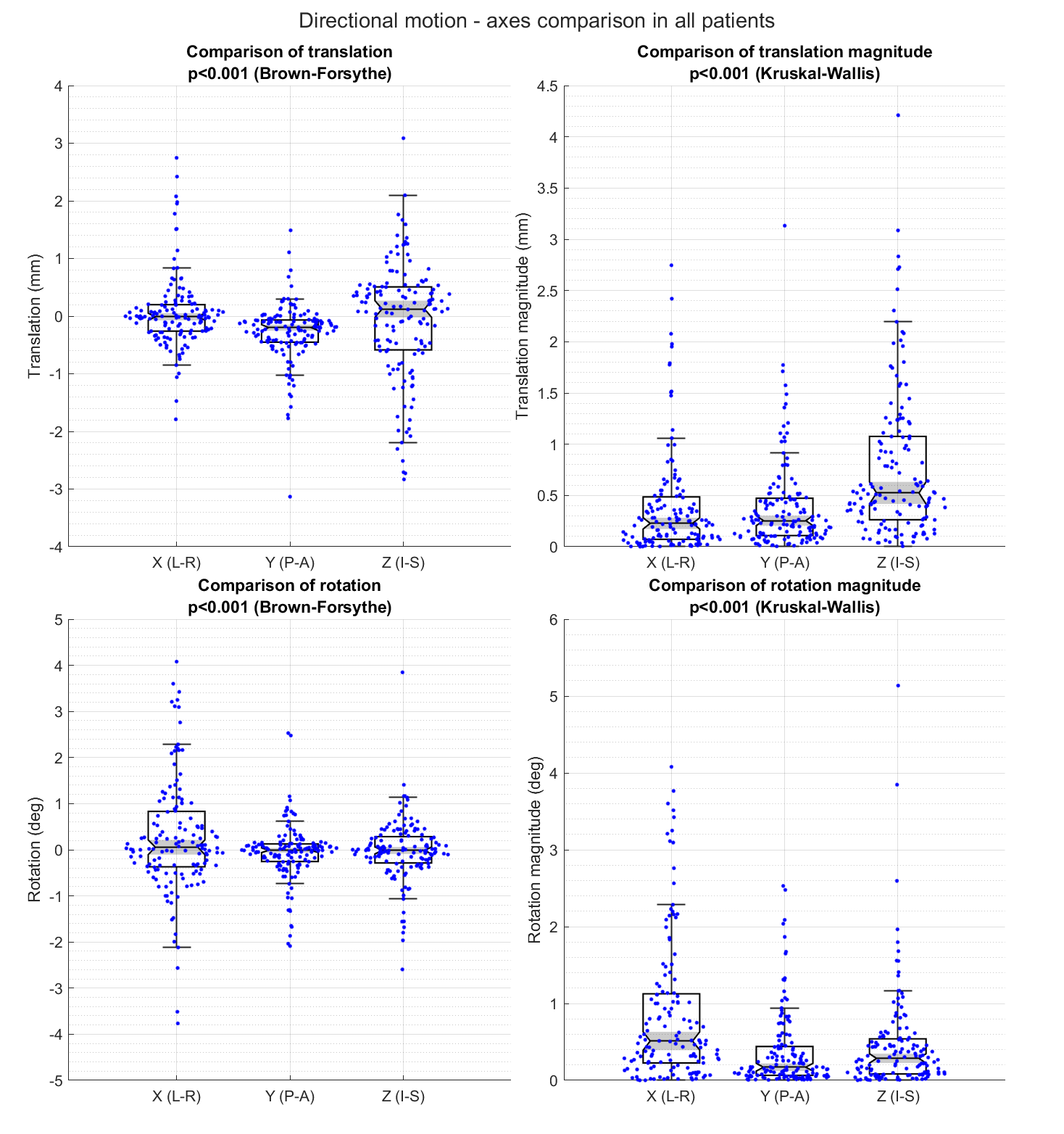

Figure S4. Translation and rotation are shown for the left–right (X), posterior–anterior (Y), and inferior–superior (Z) axes. Directional motion is shown in the left panels and absolute motion magnitude in the right panels. Statistical comparisons across axes were performed using the Brown–Forsythe test for directional motion and the Kruskal–Wallis test for absolute motion magnitude. Individual examinations are shown as points.

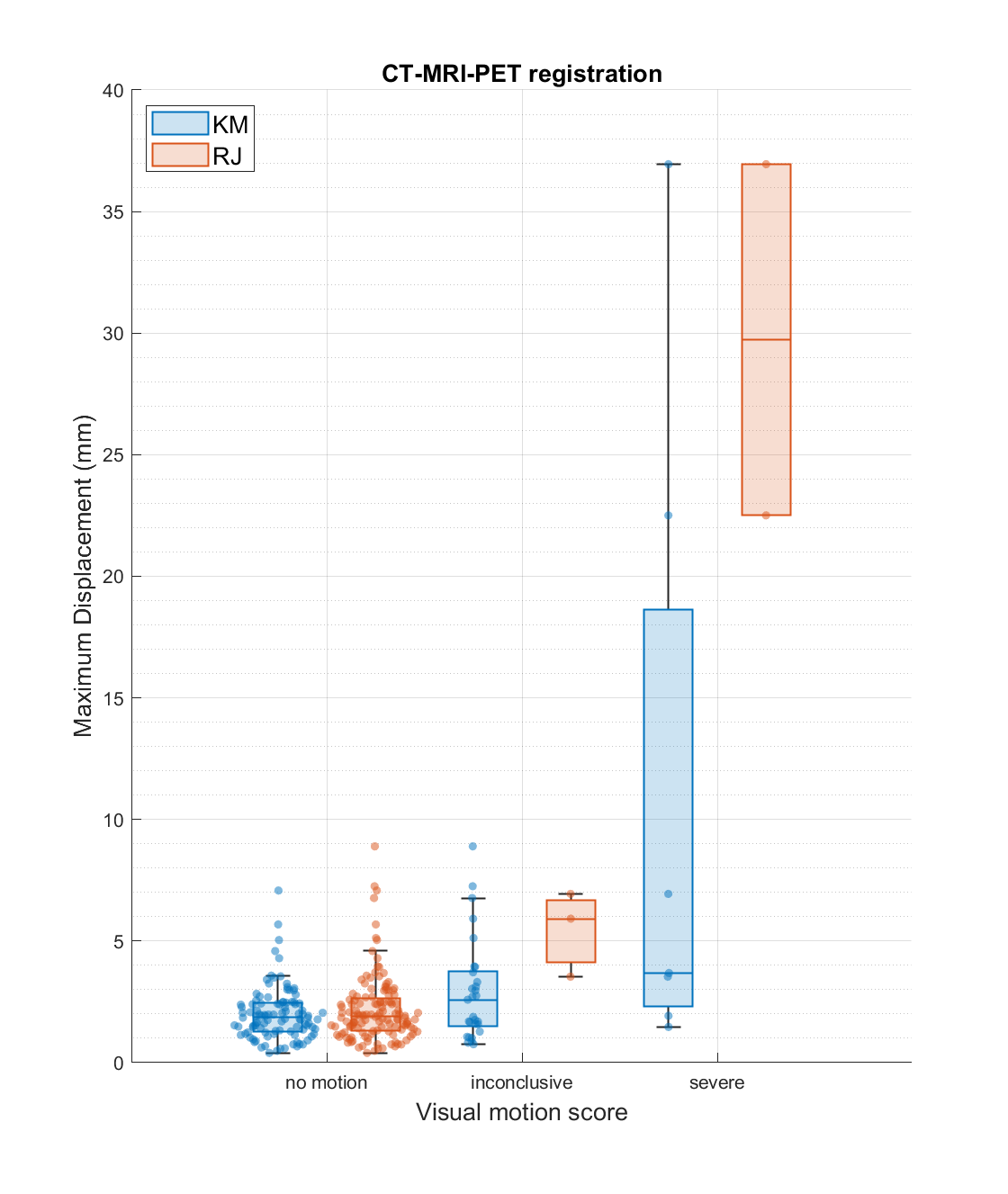

Figure S5. Comparison of visual CT–PET_1_ motion scores between readers and their relationship with displacement. Displacement between the attenuation CT and first PET frame is shown according to the visual motion scores assigned independently by the two readers. Individual examinations are shown as points; box plots indicate the median and interquartile range.
