## Supplementary figures and images for "Quantification of patient motion in FDG-PET/CT brain imaging"

### Supplementary File 2

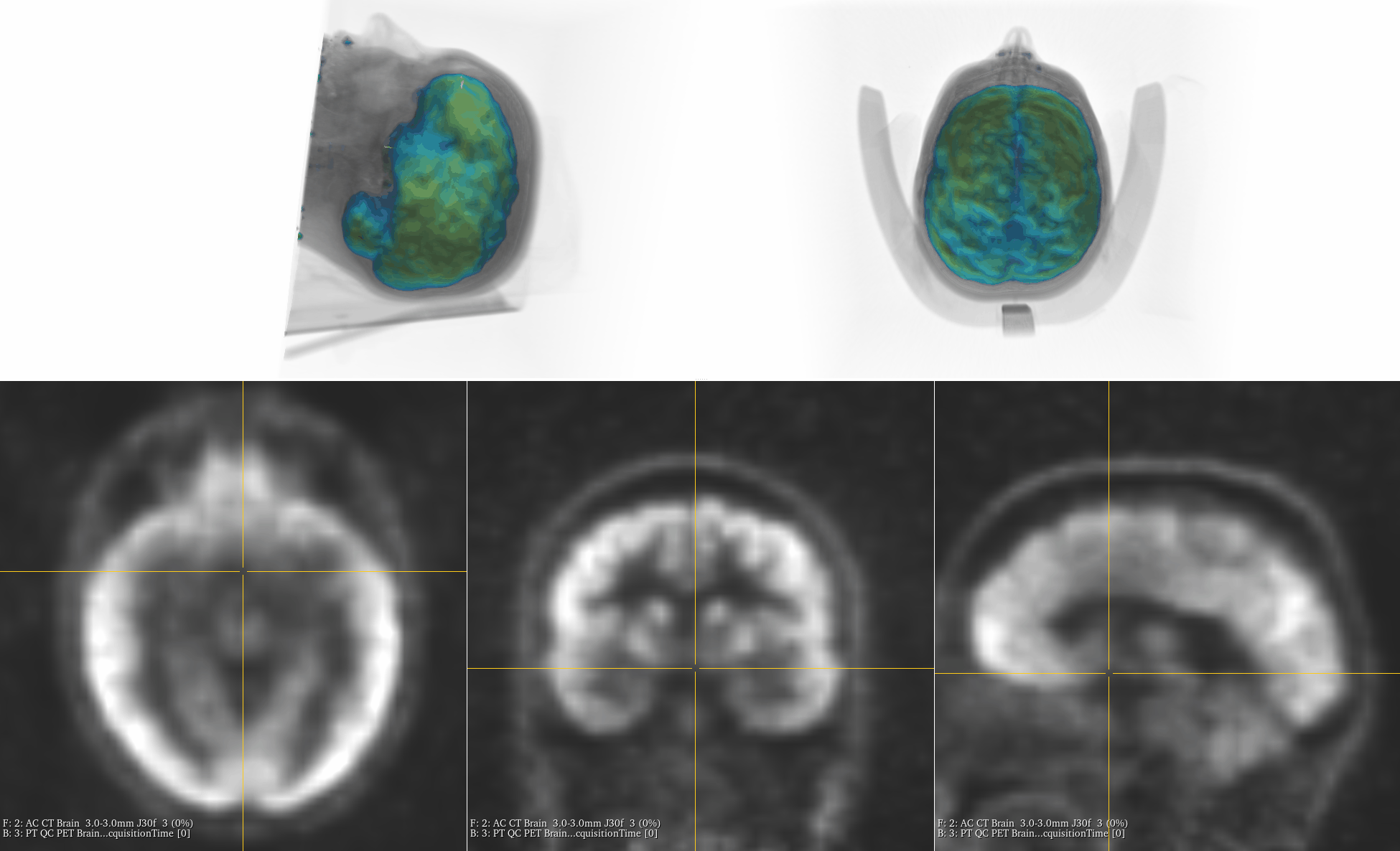
